# Price Per Standard Drink and Alcohol-Related Outcomes Among Vulnerable Groups in British Columbia: Findings from the Consumption, Harms, Expenditures and Alcohol Prices Study

**DOI:** 10.64898/2026.03.05.26347738

**Authors:** James Clay, Keegan Lawrence, Priya K. Johal, Adam Sherk, Elizabeth K. Farkouh, Tim Stockwell, Timothy Naimi

**Affiliations:** Canadian Institute for Substance Use Research (CISUR), University of Victoria, Victoria, British Columbia, Canada; Department of Community Health and Epidemiology, Dalhousie University, Halifax, Nova Scotia, Canada; Psychology Department, University of Victoria, British Columbia, Canada; Canadian Centre on Substance Use and Addiction, Ottawa, Ontario, Canada; School of Public Health and Social Policy, University of Victoria, Canada; Brigham & Women’s Hospital, Harvard Medical School, Boston, Massachusetts, USA; School of Medical Sciences, University of Victoria, Canada

**Keywords:** Alcohol Drinking, Price Per Standard Drink, Minimum Unit Pricing, Health Equity, Vulnerable Populations, Canada

## Abstract

**Objective:** Minimum unit pricing (MUP) reduces use of cheap, high strength alcoholic beverages that drive harm, yet concerns remain about inequitable effects for structurally vulnerable groups. As part of the *Consumption, Harms, Expenditures and Alcohol Prices (CHEAP)* study, we linked individual-level, product-specific alcohol consumption survey data with provincial retail price data to estimate prices per standard drink (PPSD) and examine their association with alcohol-related outcomes across sociodemographic groups.

**Method:** A cross-sectional survey of people who consumed alcohol in the past week in British Columbia, Canada, was linked to provincial product-level alcohol sales data. The population weighted sample included 1,217 adults ≥ 19 years (716 men; mean age 49.34, SD 16.98). Participants reported product-specific consumption, which was matched to retail prices to calculate individual-level PPSD. Survey weighted quasibinomial models examined associations between PPSD and three outcomes: (1) causing harm to self or others in the past year, (2) scoring ≥ 8 on the Alcohol Use Disorder Identification Test, and (3) consuming ≥ 15 standard drinks per week. Analyses were stratified by income, education, subjective social status, and race/ethnicity.

**Results:** Lower price per standard drink was associated with higher odds of harm (OR 3.05, 95% CI 1.25–7.40) and an AUDIT score ≥ 8 (OR 2.34, 95% CI 1.37–3.99). Associations were generally stronger among structurally disadvantaged groups, including low-income and Indigenous participants.

**Conclusions:** Lower alcohol prices are linked to risky alcohol use, with the strongest effects among structurally disadvantaged groups. MUP is likely to reduce this risk and promote health equity.

## Introduction

Alcohol is a widely used, addictive substance that contributes to substantial health and social harm (Babor et al., 2023). Alcohol use contributes to more than 200 disease and injury conditions and imposes major economic burdens through violence, healthcare utilisation, lost productivity, and premature mortality (World Health Organization, 2024).

It is widely known that greater volumes of alcohol consumption are the primary driver of alcohol-related harm (Rehm et al., 2010). Consequently, policies that influence the affordability of alcohol are consistently identified as the most effective mechanisms for reducing alcohol consumption and therefore related harm (World Health Organization, 2019). Among these, minimum unit pricing (MUP) which establishes a minimum price per fixed amount of ethanol, is considered the most effective affordability policy as it targets cheap, often high strength alcohol typically consumed by individuals at higher risk of harm, including those who engage in risky drinking, are dependent, or are otherwise vulnerable (Anderson et al., 2024; Black et al., 2011; Boniface et al., 2017; Chick et al., 2016).

Alcohol affordability is also an important equity concern. Individuals from low-income or otherwise marginalised groups often drink less than their more advantaged counterparts yet experience greater harm per drink consumed; a pattern known as the alcohol-harm paradox (Boyd et al., 2022). This disproportionate burden reflects cumulative disadvantage, second-hand harms, and structural inequities that heighten vulnerability (Rosen et al., 2024). Research also links heavy drinking to worsening poverty and reduced employment (Johansson et al., 2007; Jørgensen et al., 2019). Reducing consumption through pricing therefore holds potential for improving health equity, with global evidence showing disproportionate benefits for structurally vulnerable groups when alcohol prices increase (Clay et al., 2025).

BC has a longstanding history of using price-based alcohol policy, with population-level analyses demonstrating that a 10% increase in the minimum price was associated with reductions in alcohol sales, hospital admissions, and mortality (Stockwell et al., 2012, 2013; Zhao et al., 2013). However, BC’s current graduated pricing system is not fully aligned with ethanol content and is not consistently indexed to inflation, and public health experts argue the province is overdue for a true MUP (Health Officers’ Council of British Columbia, 2025; Naimi et al., 2023).

Despite strong evidence for the effectiveness and equity benefits of affordability-based interventions, major gaps remain in understanding how alcohol price relates to consumption patterns and harm among structurally vulnerable groups. Existing research has relied largely on aggregate sales data or modelling assumptions, meaning that the relationship between product-level price and individual-level consumption has not been directly measured. Policymakers therefore lack empirical evidence on the types of products consumed by different population groups, the price per standard drink (PPSD) distribution associated with those products, and - crucially - how exposure to cheap, high-risk alcohol varies across structurally vulnerable groups who may benefit most from price-based interventions. Yet no previous study has linked population-representative, product-level survey data to verified retail pricing information in a quasi-monopoly setting such as BC, where centrally regulated prices minimise measurement error inherent in self-reported expenditure data.

We therefore used data from the **C**onsumption, **H**arms, **E**xpenditures and **A**lcohol **P**rices (CHEAP) study, which surveyed individuals across BC who consumed alcohol in the past week and linked their reported product-specific consumption to provincial price data to calculate individual-level PPSD. Using population weights, we then examined how PPSD varied across structurally vulnerable groups and assessed its association with markers of harm, hazardous drinking, and high-volume consumption. This analysis provides new insight into alcohol affordability across structurally vulnerable groups, offers essential evidence to guide more equitable pricing policies, and generates high-resolution inputs that can improve the accuracy of future policy models evaluating alcohol tax and pricing reforms.

## Methods

### Study design

This cross-sectional study surveyed adults in BC who had consumed alcohol in the past seven days. Data were collected using REDCap (Harris et al., 2009, 2019). Survey responses were linked to product-specific retail price data to estimate the PPSD for each reported product and the PPSD profile among each respondent. We examined associations between lower PPSD and three outcomes: (1) self-reported alcohol-related harms to self or others in the past year (yes vs no); (2) Alcohol Use Disorders Identification Test (AUDIT) score ≥ 8 (yes vs no); and (3) consuming ≥ 15 standard drinks in the past week (yes vs no). Analyses were stratified by income, education, subjective social status, and race/ethnicity. The study protocol approved by the University of Victoria Human Research Ethics Board (#24-0235).

### Recruitment and sample

Participants were recruited between 5 and 19 March 2025 through *Leger Opinion* (LEO; a Canadian market research and survey company). Eligible respondents were BC residents aged 19 years or older (the provincial legal drinking age) who reported consuming alcohol in the past seven days.

Leger used proprietary sampling and quota procedures to recruit a sample of individuals endorsing past-week alcohol use that was broadly representative by age, gender, and geographic region, based on Statistics Canada census data. Invitations were distributed via the LEO platform and mobile app.

The average survey completion time was 12.23 minutes (SD = 40.01). Of the 3,565 individuals who initiated the survey, 1,774 were screened out because they did not consume alcohol in the past seven days or they did not reside in BC. Among the 1,791 eligible respondents, 1,221 (68.2%) completed all relevant items and were included in the analytic sample. A comparison of the analytic sample versus eligible non-completers across key sociodemographic characteristics is provided in Appendix A.

## Measures

### Sociodemographic characteristics

Participants reported their age, sex assigned at birth, racial or ethnic background, highest level of educational attainment, gross household income, geographic region (based on postal code forward sortation area), housing stability concerns, and subjective social status using the social ladder method (Adler et al., 2000).

### Alcohol consumption

Alcohol consumption was assessed using items capturing frequency, quantity, binge drinking, beverage types, and product-specific use. Participants reported the number of days they consumed alcohol in the past 30 days and, on drinking days, the approximate number of standard drinks consumed. A Canadian standard drink was defined for participants using typical amounts for beer, wine, spirits, and other beverages.

Binge drinking was assessed using sex-specific thresholds: men reported occasions of five or more drinks, and women reported occasions of four or more drinks, in the past 30 days and past seven days. Respondents also reported the maximum number of drinks consumed on a single occasion in the past 30 days.

To capture product-specific use, participants selected which alcohol products they had consumed in the past seven days across beverage categories (beer, wine, spirits, refreshment beverages; Appendix B). Product lists were constructed from retail sales data and covered items accounting for the majority of market share in 2022 in each category. For each selected product, participants reported the number of standard drinks consumed.

Weekly alcohol intake was calculated by summing product-level standard drinks consumed in the past seven days and then categorised into four groups: low (1–2 standard drinks per week), medium (3–7), high (8–14), and highest (≥ 15). These categories align approximately with Canada’s Guidance on Alcohol and Health (Paradis et al., 2023), with a further subdivision above seven drinks per week (Paradis et al., 2023).

Participants also reported the total amount of money spent on alcohol in the past seven days and estimated the percentage of drinks consumed in on-premise settings (e.g., at bars, restaurants, clubs). Finally, participants rated agreement with the statement, *“Price is an important factor when I decide which alcohol brand to purchase”*, on a five-point Likert scale (1 = *“Strongly disagree”* to 5 = *“Strongly agree”*).

### Alcohol-related harms

Alcohol-related harms were measured with three separate self-report items asking whether there had ever been a time when: (1) the participant’s own alcohol use had a harmful effect on their life; (2) their drinking had a harmful effect on someone else’s life; and (3) someone else’s drinking had a harmful effect on the participant’s life. Each item included examples (health consequences such as violence or injuries; family, social, financial, employment, housing; and legal problems) and used a four-point response scale: 1 = *“Never”*, 2 = *“Yes, in the past year”*, 3 = *“Yes, but not in the past year”*, and 4 = *“I don’t know”*. The alcohol-related harm outcome in this study was defined as causing harm to self or others in the past year (i.e., if they answered *“Yes, in the past year”* to either item asking whether their own drinking harmed their life or someone else’s life).

### Alcohol Use Disorders Identification Test

The ten-item Alcohol Use Disorders Identification Test (AUDIT) is a screening instrument for risk of possible alcohol use disorder (Babor et al., 2001). Each item is scored from 0 to 4, yielding a total score from 0 to 40. For this analysis, respondents were classified as engaging in risky drinking if their total AUDIT score was ≥ 8 (medium risk or higher) (Babor et al., 2001).

### Product-level survey-sales data linkage

Survey data on product-specific use were linked to external sales data obtained from the British Columbia Liquor Distribution Branch (LDB), which regulates distribution, importation, and retail sale of beverage alcohol in the province. The LDB dataset included product name, stock-keeping unit (SKU), beverage type, alcohol by volume (ABV), container size, wholesale price, and retail price for products sold in government-operated BC Liquor Stores.

Products reported by respondents were matched to corresponding SKUs using automated and manual review to harmonise naming conventions and product variants. Where multiple SKUs existed for a given product, the format with the largest market share (in Canadian standard drinks) was used.

If a product was not listed in the 2024 LDB dataset, the most recent available retail price from 2023 or 2022 was used. For products not sold in government stores (e.g., winery-only wines), where retail prices were unavailable, retail prices were imputed using the wholesale price and the mean mark-up ratio for that beverage category (beer, wine, spirits, refreshment beverages) and year. The mark-up ratio was calculated from products with known wholesale and retail prices as (retail price − wholesale price) / wholesale price, and averaged within each category (beer, wine, spirits, refreshment beverages) and year.

The number of standard drinks per container was calculated by multiplying container volume by ABV and dividing by 17.05 mL, the volume of pure ethanol in a Canadian standard drink (13.45 g). PPSD for each product was then calculated as retail price divided by the number of standard drinks.

These product-level PPSD values were merged with respondents reported consumption data to generate individual-level price estimates. Average PPSD was calculated as a weighted mean, where each product’s PPSD was weighted by the number of standard drinks consumed. Total alcohol expenditure in the past seven days was computed by multiplying the number of standard drinks for each product by its PPSD and summing across all products.

### Survey weighting

Post-stratification weights were applied to improve representativeness of people who drank in the past week in BC. Population benchmarks were derived from the 2019 Canadian Alcohol and Drugs Survey (Statistics Canada, 2021) using marginal distributions for age group (19–25, 26–44, 45–64, 65+), sex (male, female), and household income (prefer not to answer, < CAD 40,000, CAD 40,000–79,999, ≥ CAD 80,000).

Raking (iterative proportional fitting) was implemented using the survey package in R to align sample margins with population distributions (Deville & and Särndal, 1992). Weights were trimmed at the 2.5th and 97.5th percentiles to limit extreme values and then normalized to sum to the total sample size. Trimmed weights were used for estimating means and totals, and normalized weights were used in regression models.

### Statistical analysis

Four outliers were removed *a priori* (three reporting > 150 standard drinks/week; one reporting PPSD > CAD 15), leaving 1,217 respondents. Descriptive statistics (means, standard deviations and proportions) were calculated for variables of interest and select demographic variables.

Survey-weighted logistic regression models were estimated using the svyglm function in the survey package for R, specifying a quasibinomial family to account for overdispersion. Results are presented as odds ratios (ORs) with 95 per cent confidence intervals (CIs). PPSD was log-transformed prior to modelling to improve distributional properties.

Socioeconomic variables (income, education, and social ladder) were each collapsed into three categories: low (income: CAD $30,000–59,999; education: high school or less; social ladder: 0–3), medium (income: CAD $60,000–124,999; education: college or bachelor’s degree; ladder: 4–7), and high (income: CAD $125,000+; education: master’s or doctoral degree; ladder: 8–10). Ethnicity was collapsed into two sets of binary indicators (White versus non-White and Indigenous versus non-Indigenous).

We first estimated bivariate models regressing each outcome on PPSD (Model 1), then sequentially and cumulatively adjusted for possible covariates: age (continuous; Model 2), sex (men vs. women; Model 3), ethnicity (White vs. non-White; Model 4), and income (low, medium, high; Model 5). Responses coded as “prefer not to answer” were retained in these models to preserve sample size. Finally, stratified models were estimated for each socioeconomic and race/ethnicity subgroup, adjusting only for age and sex to reduce overfitting and preserve model stability in smaller subgroups.

Analyses were exploratory and not pre-registered. All analyses were conducted in R (version 4.3.2). A two-tailed p-value < 0.05 was considered statistically significant.

## Results

### Sample characteristics

Table 1 presents descriptive statistics for the sociodemographic characteristics of the sample and key study variables. Respondents had a mean age of 49.34 years (SD = 16.98), and the weighted sample was predominantly White (68.09%), with 22.72% identifying as Asian and smaller proportions as Indigenous, Hispanic/Latin, Black, or other ethnicities.

**Table 1.** Sociodemographic characteristics of the sample and descriptive statistics for study variables.

| Variable | Combined (SD) | Women (SD) | Men (SD) |
| --- | --- | --- | --- |
| N | 1217 | 501 | 716 |
| Age | 49.34 (16.98) | 46.90 (17.09) | 51.16 (16.68) |
| Ethnicity |  |  |  |
| Arab or Middle Eastern | 1.18% | 0.38% | 1.79% |
| Asian | 22.72% | 20.34% | 24.51% |
| Black | 0.97% | 0.35% | 1.43% |
| Hispanic or Latin | 1.38% | 2.32% | 0.67% |
| Indigenous | 1.88% | 2.48% | 1.44% |
| Other | 0.79% | 1.64% | 0.16% |
| White | 68.09% | 70.34% | 66.41% |
| Prefer not to answer | 2.98% | 2.16% | 3.59% |
| Education |  |  |  |
| Some high school | 0.81% | 0.44% | 1.08% |
| High school or GED equivalent | 13.92% | 14.81% | 13.25% |
| College diploma or equivalent | 30.84% | 35.37% | 27.45% |
| Bachelors | 36.81% | 37.26% | 36.48% |
| Masters | 14.83% | 11.13% | 17.60% |
| Doctoral | 2.20% | 0.78% | 3.27% |
| Prefer not to answer | 0.59% | 0.22% | 0.87% |
| Income |  |  |  |
| < \$30,000 | 3.94% | 4.45% | 3.55% |
| \$30,000-\$59,999 | 8.52% | 10.02% | 7.40% |
| \$60,000-\$99,999 | 23.94% | 24.78% | 23.31% |
| \$100,000-\$124,999 | 21.49% | 18.30% | 23.88% |
| \$125,000-\$199,999 | 22.98% | 21.25% | 24.28% |
| \$200,000 or more | 8.96% | 6.89% | 10.51% |
| Prefer not to answer | 10.17% | 14.30% | 7.07% |
| Social ladder | 6.20 (1.72) | 6.06 (1.70) | 6.31 (1.73) |
| Worry about housing = Yes | 12.17% | 11.79% | 12.45% |
| Self-report drinks per week | 8.21 (10.79) | 7.52 (10.57) | 8.73 (10.94) |
| Self-report on-premise consumption | 25.46% | 26.21% | 24.91% |
| Binge frequency (past 7 days) | 0.71 (1.47) | 0.76 (1.57) | 0.68 (1.40) |
| AUDIT | 5.94 (6.03) | 5.48 (5.81) | 6.29 (6.16) |
| AUDIT categories |  |  |  |
| Low (0-7 points) | 76.57% | 78.85% | 74.85% |
| Medium (8-15 points) | 15.14% | 13.47% | 16.39% |
| High (16-19 points) | 3.01% | 2.97% | 3.04% |
| Possible AUD (20+ points) | 4.93% | 4.13% | 5.53% |
| Prefer not to answer | 0.36% | 0.59% | 0.19% |
| Drinking group, past 7 days |  |  |  |
| Low (1-2 drinks) | 31.06% | 36.38% | 27.08% |
| Medium (3-7 drinks) | 34.26% | 32.25% | 35.77% |
| High (8-14 drinks) | 20.29% | 18.71% | 21.47% |
| Highest (15+ drinks) | 14.38% | 12.65% | 15.68% |
| Price importance | 3.65 (1.11) | 3.71 (1.06) | 3.61 (1.14) |
| Weekly alcohol expenditure (CAD) | 19.75 (26.91) | 18.92 (26.32) | 20.36 (27.35) |
| Price per standard drink (CAD) | 2.38 (1.08) | 2.54 (1.11) | 2.27 (1.05) |
| Harm to self (own drinking) |  |  |  |
| Never | 64.63% | 66.32% | 63.36% |
| Yes, in the past year | 16.58% | 15.63% | 17.29% |
| Yes, but not in the past year | 16.50% | 15.63% | 17.15% |
| I don't know | 2.29% | 2.43% | 2.19% |
| Harm to others (own drinking) |  |  |  |
| Never | 72.77% | 73.13% | 72.49% |
| Yes, in the past year | 10.22% | 9.95% | 10.43% |
| Yes, but not in the past year | 13.47% | 13.23% | 13.65% |
| I don't know | 3.54% | 3.69% | 3.44% |
| Harm to self (others' drinking) |  |  |  |
| Never | 60.00% | 52.90% | 65.31% |
| Yes, in the past year | 13.16% | 14.49% | 12.16% |
| Yes, but not in the past year | 23.32% | 28.59% | 19.37% |
| I don't know | 3.52% | 4.02% | 3.15% |
| First-hand harm (past year) | 6.99% | 6.91% | 7.05% |

*Note.* Means, standard deviations, and percentages reflect weighted statistics. Binge drinking was defined as consuming four or more alcoholic beverages on a single occasion for women, and five or more for men. AUDIT = Alcohol Use Disorders Identification Test. AUD = Alcohol Use Disorder. Past-year alcohol harm was defined as participants responding “*Yes, in the past year*” to either (a) experiencing harmful effects from their own drinking or (b) their drinking causing harm to someone else. Harm to self (own drinking) = harmful effects from one’s own drinking; Harm to others (own drinking) = one’s own drinking causing harm to someone else; Harm to self (others’ drinking) = harmful effects from someone else’s drinking. Past-year first-hand harm was defined as participants responding ‘*Yes, in the past year*’ to either (a) harm to self (own drinking) or (b) harm to others (own drinking).

On average during the past 7 days, participants consumed 8.21 drinks (SD = 10.79) and spent CAD $19.75 (SD = 26.91) on alcohol. Mean PPSD was CAD $2.38 (SD = 1.08), with a positively skewed distribution and most values below CAD 4.00 (Figure 1). Respondents reported consuming an average of 25.46% of drinks on-premise, with 57.74% reporting at least some on-premise alcohol consumption (Appendix C). 31.06% were classified as engaging in low-risk drinking, 34.26% medium-risk, 20.29% high-risk, and 14.38% highest-risk. AUDIT scores classified 76.57% of respondents as low-, 15.14% as medium-, 3.01% as high-risk, and 4.93% as possible alcohol use disorder.

**Figure 1.**
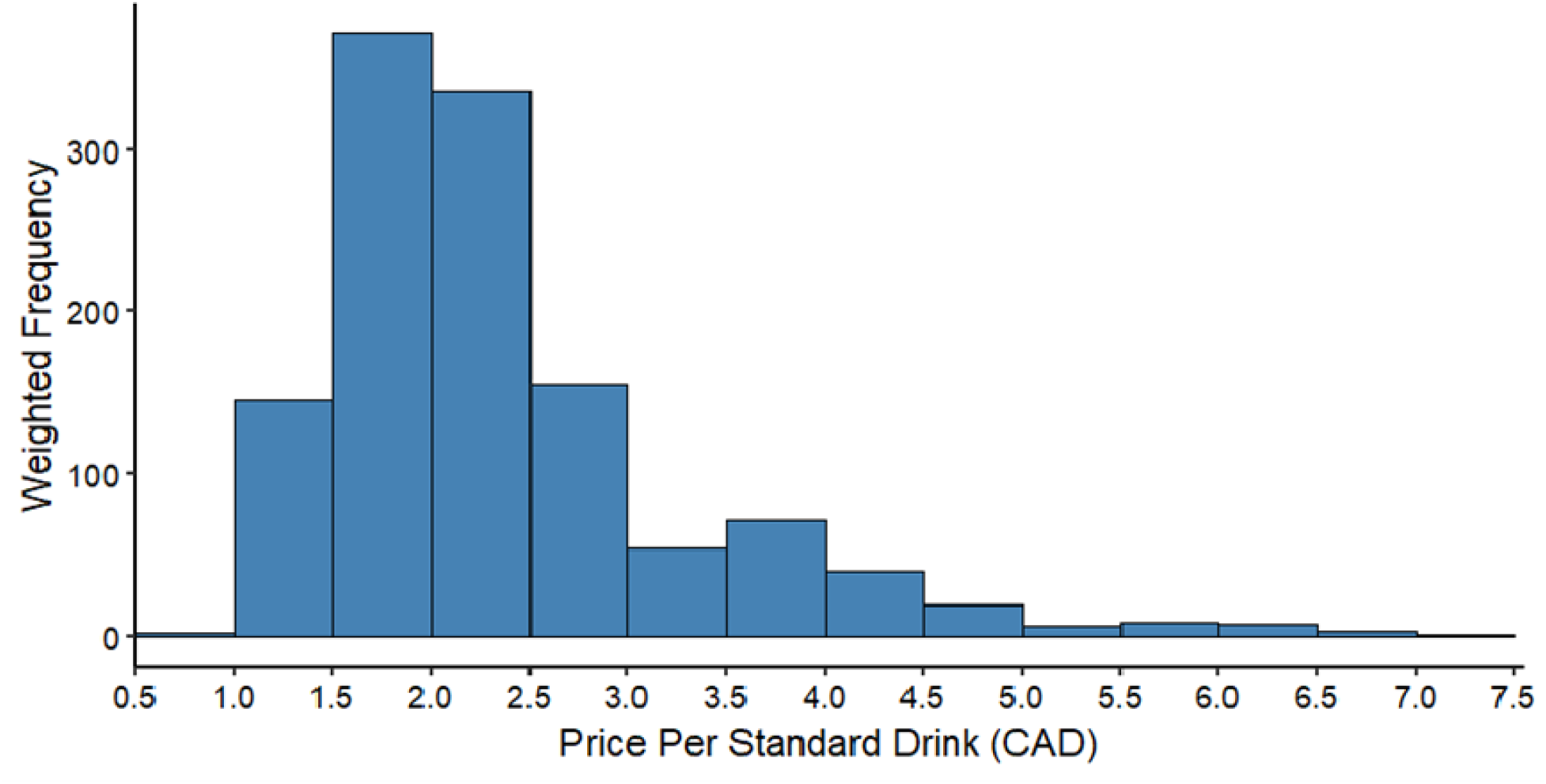
Histogram of weighted price per standard drink (PPSD) among persons who consumed alcohol in the past seven days (*n* = 1215). Note. The x-axis is truncated at $7.50 CAD to improve readability. Values above this threshold were excluded from the figure but retained in all analyses. This truncation removed approximately 0.16% of observations. The maximum observed mean PPSD value was $14.53 CAD. The histogram display survey-weighted frequencies, with a bin width of $0.50 CAD.

### Association between PPSD and alcohol-related outcomes

Lower mean log(PPSD) was generally related to greater odds of adverse alcohol-related outcomes (Table 2). In unadjusted models, each CAD $1.00 decrease in log(PPSD) was associated with higher odds of an AUDIT score ≥ 8 (OR = 1.89, 95% CI: 1.19–3.00, *p* = 0.007), while associations with past-year harms (OR = 2.04, 95% CI: 0.99–4.23, *p* = 0.054) and consuming ≥ 15 drinks per week (OR = 1.58, 95% CI: 0.96–2.61, *p* = 0.071) were associated with higher odds but not statistically significant.

**Table 2.** Adjusted odds ratios for the association between lower price per standard drink and alcohol-related outcomes.

| **Model** | **Harm**<br>**(95% CI)**<br>***n* = 1217** | **AUDIT ≥ 8**<br>**(95% CI)**<br>***n* = 1212** | **≥ 15 Drinks**<br>**(95% CI)**<br>***n* = 1217** |
| --- | --- | --- | --- |
| Model 1 | 2.04 (0.99-4.23) | **1.89 (1.19-3.00)** | 1.58 (0.96-2.61) |
| Model 2 | **2.93 (1.26-6.79)** | **2.67 (1.58-4.51)** | 1.40 (0.87-2.26) |
| Model 3 | **2.88 (1.2-6.87)** | **2.53 (1.49-4.31)** | 1.36 (0.83-2.23) |
| Model 4 | **3.02 (1.27-7.17)** | **2.54 (1.50-4.32)** | 1.34 (0.82-2.19) |
| Model 5 | **3.05 (1.25-7.40)** | **2.34 (1.37-3.99)** | 1.31 (0.80-2.16) |

Note. Values represent odds ratios (ORs) and 95% confidence intervals from logistic regression models predicting (1) self-reported past-year alcohol harms vs. not; (2) reporting an AUDIT score ≥ 8 vs. not; and (3) ≥15 drinks per week. Odds refer to the odds of an outcome per \$1.00 decrease in PPSD. AUDIT = Alcohol Use Disorders Identification Test. Harm = experiencing a first-hand harm in the past year, defined as: a participant responded “*Yes, in the past year*” to either (a) experiencing harmful effects from their own drinking or (b) their drinking causing harm to someone else. Boldface indicates statistical significance ( $p < 0.05$ ).
Model 1 = Bivariate
Model 2 = Model 1 + age
Model 3 = Model 2 + sex
Model 4 = Model 3 + ethnicity
Model 5 = Model 4 + income

After sequential adjustment for sociodemographic covariates, associations became stronger. In fully adjusted models, lower log(PPSD) was associated with higher odds of past-year alcohol-related harms (OR = 3.05, 95% CI: 1.25–7.40, *p* = 0.014) and an AUDIT score ≥ 8 8 (OR = 2.34, 95% CI: 1.37– 3.99, *p* = 0.002). The association with heavy weekly consumption showed similar patterns but was non-significant (OR = 1.31, 95% CI: 0.80–2.16, *p* = 0.286).

### Socioeconomic variation

Patterns of affordability and risk varied across socioeconomic groups (Tables 3 and 4). Respondents with lower income, lower educational attainment, or lower subjective social status generally purchased cheaper alcohol and exhibited higher levels of risky outcomes.

**Table 3.**
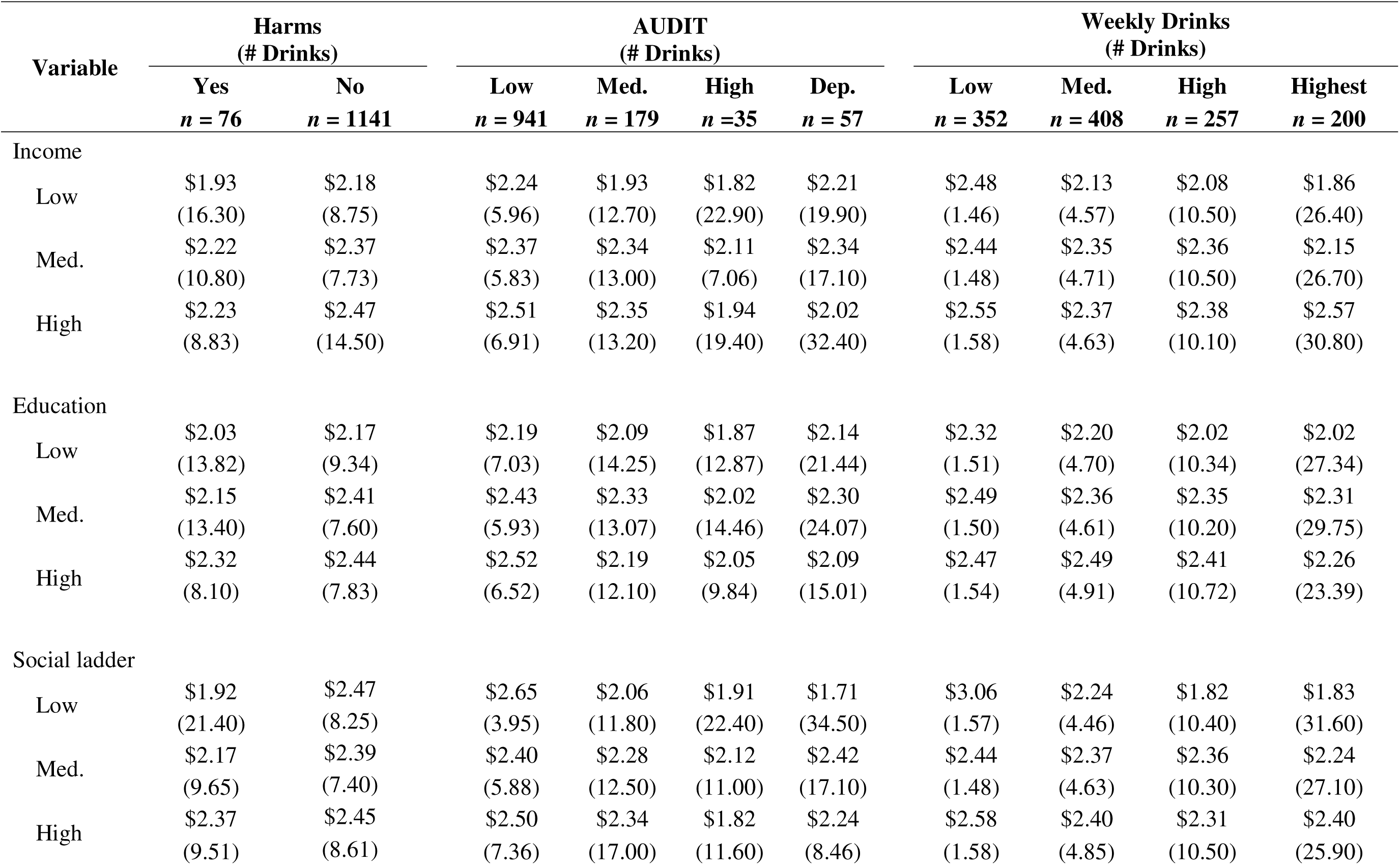
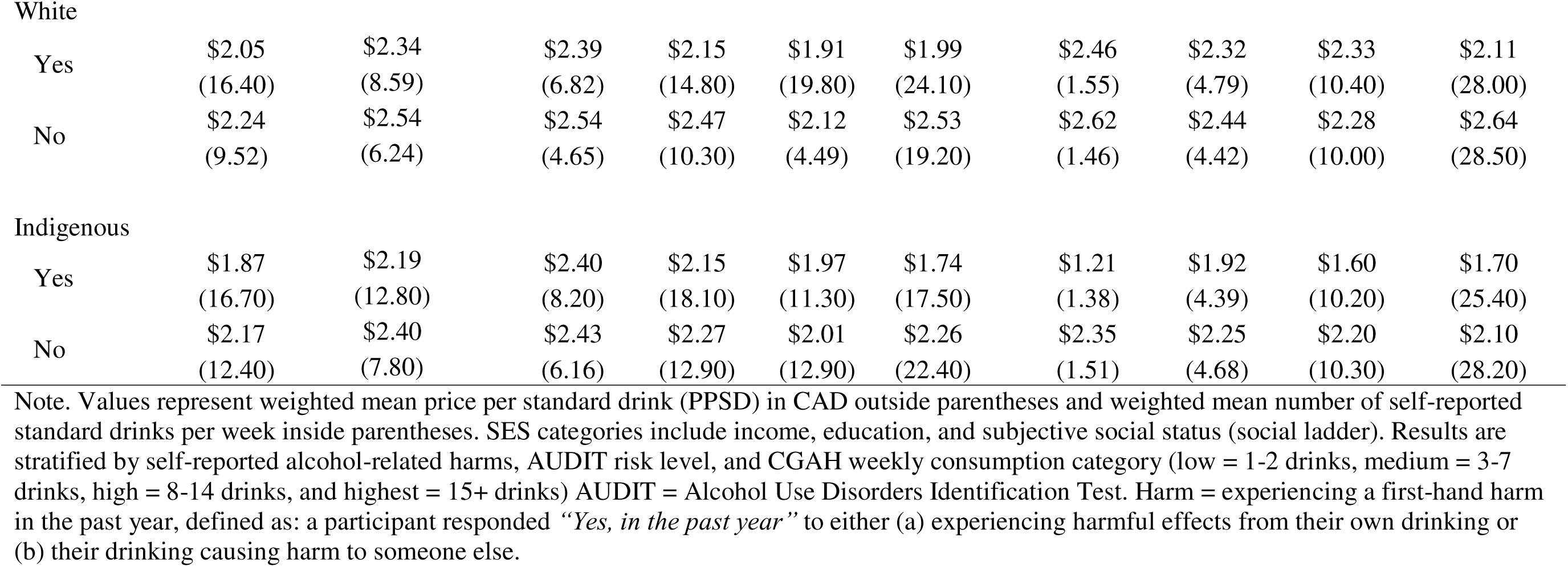
Mean price per standard drink and number of drinks consumed among sociodemographic groups by self-reported harm, AUDIT risk level, and alcohol consumption.

| Variable | Harms<br>(# Drinks) |  | AUDIT<br>(# Drinks) |  |  |  | Weekly Drinks<br>(# Drinks) |  |  |  |
| --- | --- | --- | --- | --- | --- | --- | --- | --- | --- | --- |
|  | Yes<br>*n* = 76 | No<br>*n* = 1141 | Low<br>*n* = 941 | Med.<br>*n* = 179 | High<br>*n* = 35 | Dep.<br>*n* = 57 | Low<br>*n* = 352 | Med.<br>*n* = 408 | High<br>*n* = 257 | Highest<br>*n* = 200 |
| Income |  |  |  |  |  |  |  |  |  |  |
| Low | \$1.93<br>(16.30) | \$2.18<br>(8.75) | \$2.24<br>(5.96) | \$1.93<br>(12.70) | \$1.82<br>(22.90) | \$2.21<br>(19.90) | \$2.48<br>(1.46) | \$2.13<br>(4.57) | \$2.08<br>(10.50) | \$1.86<br>(26.40) |
| Med. | \$2.22<br>(10.80) | \$2.37<br>(7.73) | \$2.37<br>(5.83) | \$2.34<br>(13.00) | \$2.11<br>(7.06) | \$2.34<br>(17.10) | \$2.44<br>(1.48) | \$2.35<br>(4.71) | \$2.36<br>(10.50) | \$2.15<br>(26.70) |
| High | \$2.23<br>(8.83) | \$2.47<br>(14.50) | \$2.51<br>(6.91) | \$2.35<br>(13.20) | \$1.94<br>(19.40) | \$2.02<br>(32.40) | \$2.55<br>(1.58) | \$2.37<br>(4.63) | \$2.38<br>(10.10) | \$2.57<br>(30.80) |
| Education |  |  |  |  |  |  |  |  |  |  |
| Low | \$2.03<br>(13.82) | \$2.17<br>(9.34) | \$2.19<br>(7.03) | \$2.09<br>(14.25) | \$1.87<br>(12.87) | \$2.14<br>(21.44) | \$2.32<br>(1.51) | \$2.20<br>(4.70) | \$2.02<br>(10.34) | \$2.02<br>(27.34) |
| Med. | \$2.15<br>(13.40) | \$2.41<br>(7.60) | \$2.43<br>(5.93) | \$2.33<br>(13.07) | \$2.02<br>(14.46) | \$2.30<br>(24.07) | \$2.49<br>(1.50) | \$2.36<br>(4.61) | \$2.35<br>(10.20) | \$2.31<br>(29.75) |
| High | \$2.32<br>(8.10) | \$2.44<br>(7.83) | \$2.52<br>(6.52) | \$2.19<br>(12.10) | \$2.05<br>(9.84) | \$2.09<br>(15.01) | \$2.47<br>(1.54) | \$2.49<br>(4.91) | \$2.41<br>(10.72) | \$2.26<br>(23.39) |
| Social ladder |  |  |  |  |  |  |  |  |  |  |
| Low | \$1.92<br>(21.40) | \$2.47<br>(8.25) | \$2.65<br>(3.95) | \$2.06<br>(11.80) | \$1.91<br>(22.40) | \$1.71<br>(34.50) | \$3.06<br>(1.57) | \$2.24<br>(4.46) | \$1.82<br>(10.40) | \$1.83<br>(31.60) |
| Med. | \$2.17<br>(9.65) | \$2.39<br>(7.40) | \$2.40<br>(5.88) | \$2.28<br>(12.50) | \$2.12<br>(11.00) | \$2.42<br>(17.10) | \$2.44<br>(1.48) | \$2.37<br>(4.63) | \$2.36<br>(10.30) | \$2.24<br>(27.10) |
| High | \$2.37<br>(9.51) | \$2.45<br>(8.61) | \$2.50<br>(7.36) | \$2.34<br>(17.00) | \$1.82<br>(11.60) | \$2.24<br>(8.46) | \$2.58<br>(1.58) | \$2.40<br>(4.85) | \$2.31<br>(10.50) | \$2.40<br>(25.90) |
| White |  |  |  |  |  |  |  |  |  |  |
| Yes | \$2.05<br>(16.40) | \$2.34<br>(8.59) | \$2.39<br>(6.82) | \$2.15<br>(14.80) | \$1.91<br>(19.80) | \$1.99<br>(24.10) | \$2.46<br>(1.55) | \$2.32<br>(4.79) | \$2.33<br>(10.40) | \$2.11<br>(28.00) |
| No | \$2.24<br>(9.52) | \$2.54<br>(6.24) | \$2.54<br>(4.65) | \$2.47<br>(10.30) | \$2.12<br>(4.49) | \$2.53<br>(19.20) | \$2.62<br>(1.46) | \$2.44<br>(4.42) | \$2.28<br>(10.00) | \$2.64<br>(28.50) |
| Indigenous |  |  |  |  |  |  |  |  |  |  |
| Yes | \$1.87<br>(16.70) | \$2.19<br>(12.80) | \$2.40<br>(8.20) | \$2.15<br>(18.10) | \$1.97<br>(11.30) | \$1.74<br>(17.50) | \$1.21<br>(1.38) | \$1.92<br>(4.39) | \$1.60<br>(10.20) | \$1.70<br>(25.40) |
| No | \$2.17<br>(12.40) | \$2.40<br>(7.80) | \$2.43<br>(6.16) | \$2.27<br>(12.90) | \$2.01<br>(12.90) | \$2.26<br>(22.40) | \$2.35<br>(1.51) | \$2.25<br>(4.68) | \$2.20<br>(10.30) | \$2.10<br>(28.20) |

Note. Values represent weighted mean price per standard drink (PPSD) in CAD outside parentheses and weighted mean number of self-reported standard drinks per week inside parentheses. SES categories include income, education, and subjective social status (social ladder). Results are stratified by self-reported alcohol-related harms, AUDIT risk level, and CGAH weekly consumption category (low = 1-2 drinks, medium = 3-7 drinks, high = 8-14 drinks, and highest = 15+ drinks) AUDIT = Alcohol Use Disorders Identification Test. Harm = experiencing a first-hand harm in the past year, defined as: a participant responded “*Yes, in the past year*” to either (a) experiencing harmful effects from their own drinking or (b) their drinking causing harm to someone else.

**Table 4.** Adjusted odds ratios for the association between lower price per standard drink and alcohol-related outcomes by socioeconomic status and race/ethnicity.

| Variable | *n* | Harm<br>(95% CI) | AUDIT ≥ 8<br>(95% CI) | ≥ 15 Drinks<br>(95% CI) |
| --- | --- | --- | --- | --- |
| Income |  |  |  |  |
| Low | 322 | 3.57 (0.84-15.20) | **4.45 (1.58-12.50)** | **3.97 (1.46-5.61)** |
| Medium | 535 | 2.13 (0.67-6.75) | 1.55 (0.75-3.21) | 1.70 (0.82-3.52) |
| High | 278 | 2.92 (0.38-22.40) | **3.02 (1.03-8.89)** | 0.60 (0.24-1.52) |
| Education |  |  |  |  |
| Low | 226 | 2.80 (0.35-22.60) | 2.82 (0.73-11.00) | 0.83 (0.49-6.81) |
| Medium | 796 | 2.63 (0.97-7.15) | **1.99 (1.02-3.85)** | 0.98 (0.55-1.74) |
| High | 189 | 3.06 (0.17-54.40) | **6.72 (1.88-24.10)** | 2.33 (0.45-12.00) |
| Social ladder |  |  |  |  |
| Low | 127 | **4.28 (1.02-17.90)** | **7.63 (1.61-36.10)** | **7.57 (1.28-44.90)** |
| Medium | 798 | **3.03 (1.03-10.60)** | 1.85 (0.98-3.50) | 1.21 (0.63-2.32) |
| High | 245 | 0.91 (0.14-6.06) | 3.82 (0.87-16.70) | 1.04 (0.40-2.69) |
| White |  |  |  |  |
| Yes | 884 | **4.94 (1.10-22.10)** | **3.75 (1.81-7.76)** | **1.86 (1.04-3.35)** |
| No | 333 | 2.17 (0.76-6.16) | 1.44 (0.67-3.09) | 0.58 (0.26-1.28) |
| Indigenous |  |  |  |  |
| Yes | 30 | 3.39 (0.35-33.10) | **22.10 (2.17-242.89)** | 0.89 (0.10-8.32) |
| No | 1187 | **2.27 (1.12-6.87)** | **2.34 (1.36-4.01)** | 1.33 (0.80-2.21) |

Note. Values represent odds ratios (ORs) and 95% confidence intervals from logistic regression models predicting: (1) self-reported past-year alcohol harms vs. not; (2) reporting an AUDIT score ≥ 8 vs. not; and (3) ≥15 drinks per week. Odds refer to the odds of an outcome per \$1.00 decrease in PPSD. AUDIT = Alcohol Use Disorders Identification Test. Harm = experiencing a first-hand harm in the past year, defined as: a participant responded “Yes, in the past year” to either (a) experiencing harmful effects from their own drinking or (b) their drinking causing harm to someone else. Models were adjusted for age and sex. Boldface indicates statistical significance ( $p < 0.05$ ).

Among low-income respondents, each unit decrease in log(PPSD) was associated with higher odds of an AUDIT score ≥ 8 (OR = 4.45, 95% CI: 1.58–12.50, *p* = 0.005) and consuming ≥ 15 drinks per week (OR = 3.97, 95% CI: 1.46–5.61, *p* = 0.007). Among respondents with lower subjective social status, lower log(PPSD) was associated with an AUDIT score ≥ 8 (OR = 7.63, 95% CI: 1.61–36.10, *p* = 0.005), harm (OR = 4.28, 95% CI: 1.02–17.90, *p* = 0.047), and heavy consumption (OR = 7.57, 95% CI: 1.28–44.90, *p* < 0.007). Associations among higher-income and higher-status groups were smaller and less consistent.

For education, associations with outcomes were concentrated among respondents with medium and high educational attainment. Lower log(PPSD) was significantly associated with higher odds of an AUDIT score ≥ 8 among medium-educated (OR = 1.99, 95% CI: 1.02–3.85, p = 0.042) and highly educated respondents (OR = 6.72, 95% CI: 1.88–24.10, p = 0.004).

### Racial and ethnic variation

Patterns of affordability and risk varied across racial and ethnic groups (Tables 3 and 4). White respondents tended to consume more alcohol at lower PPSDs than non-White respondents. Indigenous respondents, although a small subsample, reported some of the lowest PPSDs (for example, CAD 1.87 among those reporting harms).

Among White respondents, lower log(PPSD) was significantly associated with greater odds of harm (OR = 4.94, 95% CI: 1.10–22.10, p = 0.037), an AUDIT score ≥ 8 (OR = 3.75, 95% CI: 1.81–7.76, *p* < 0.001), and consuming ≥ 15 drinks per week (OR = 1.86, 95% CI: 1.04–3.35, *p* = 0.037). Among non-White respondents, associations did not reach statistical significance. For Indigenous respondents, although the sample was small (*n* = 30), models suggested very strong associations between lower log(PPSD) and AUDIT scores ≥ 8 (OR = 22.10, 95% CI: 2.17–242.89, *p* = 0.011).

## Discussion

This study examined the relationship between PPSD and alcohol-related outcomes among a population-based sample of British Columbians who had consumed alcohol in the past week, with a focus on variation across structurally vulnerable groups. Our findings show that lower PPSD is associated with higher odds of risky alcohol use. This association varies across socioeconomic and racial/ethnic subgroups, but in general, lower PPSDs were associated with increased risk in groups with structural vulnerabilities compared to those who were relatively less vulnerable.

In the full sample, lower PPSDs were associated with greater odds of alcohol-related outcomes, particularly past-year first-hand alcohol-related harms and scoring ≥ 8 on the AUDIT. Although the association with heavy weekly consumption (≥ 15 drinks) was in the same direction, it was not statistically significant in the overall sample. These findings align with prior research showing that cheaper alcohol is associated with more hazardous drinking patterns and support the core principle of MUP and other alcohol pricing policies: that increasing the price of alcohol reduces risky use (Anderson et al., 2024; Holmes, 2023; Wagenaar et al., 2009). This is consistent with PPSD capturing exposure to cheap, high-strength alcohol products rather than exerting a direct causal effect on harm.

### Socioeconomic gradients

Stratified analyses revealed socioeconomic gradients in both exposure to low-cost alcohol and its associations with outcomes. Descriptive results indicated that respondents with lower income or lower subjective social status tended to pay less per standard drink and more often reported higher consumption when experiencing harms or scoring high on the AUDIT. Regression models further showed that, among low-income respondents, lower PPSD was associated with greater odds of higher AUDIT scores and heavy consumption. Likewise, among those with low subjective social status, lower PPSD was associated with harms, higher AUDIT scores, and heavy consumption. These findings suggest that socioeconomically disadvantaged groups may experience a dual burden—greater exposure to inexpensive alcohol and greater susceptibility to its potential negative consequences. These results corroborate those from jurisdictions where MUP has been successfully implemented. For example, in Scotland, a MUP of 50p/8g ethanol resulted in an 13% reduction overall in deaths wholly attributable to alcohol, with larger effects observed in the most deprived decile (22%) versus the least deprived decile (8%) (Wyper et al., 2023).

### Education

Associations by educational attainment presented a more complex picture. Although individuals with low education reported cheaper alcohol consumption on average, statistically significant associations were observed only among medium- and highly educated respondents. Part of this pattern may reflect age composition, as students tend to be younger and drink more on that basis (Maggs & Schulenberg, 2004). A cohort effect may also contribute: older people are less likely to have attained higher education given shifts toward occupations requiring further education (Picot & Hou, 2024). In addition, some higher-educated drinkers may consume substantial quantities of inexpensive alcohol despite socioeconomic advantage, potentially due to social factors (for example, family wealth facilitating higher education) or occupational norms around drinking. Alternatively, differential effects of health literacy and public health messaging at varying levels of risk may be at play, indicating that educational attainment alone does not uniformly protect against hazardous drinking (Chisolm et al., 2014; Hasking et al., 2005). These findings suggest that pricing interventions should be complemented by tailored communication strategies that resonate across educational levels, age groups, and cohorts.

### Race and ethnicity

Racial and ethnic subgroup analyses revealed further heterogeneity. Among White respondents, lower PPSD was consistently associated with alcohol-related harms, higher AUDIT scores, and heavy consumption. Among Indigenous respondents, although the sample size was small, models suggested very strong associations between lower PPSD and higher AUDIT scores, with odds ratios substantially larger than those observed in other groups. Importantly, even the lower bound of the 95% confidence interval (2.17) indicated a meaningful effect, though the wide interval reflects limited precision. These findings are consistent with evidence from the Northern Territory of Australia, where MUP was effective in reducing alcohol-related harms in communities with large Indigenous populations (Secombe et al., 2020, 2021; Wright et al., 2021). Together, these patterns suggest that while lower alcohol prices are associated with increased risk across groups, cultural, structural, and contextual factors may influence the degree of responsiveness to price, particularly among racially marginalised populations. Moreover, these results strengthen the evidence that pricing policies can reduce alcohol-related harms and advance health equity in historically underserved communities (Clay et al., 2025).

### Strengths and limitations

This study has several strengths. First, it uses novel product-specific consumption data linked with provincial retail price data to estimate PPSD at the individual level, rather than relying on broad population-level price metrics. Second, the data was weighted to reflect population-level drinking patterns in BC, increasing generalisability. Third, stratified analyses allowed for an equity-focused lens, highlighting variation across income, education, social status, and racial/ethnic groups. Fourth, the use of survey-weighted regression with a quasibinomial specification provided robust variance estimates. Fifth, the study assessed multiple outcomes spanning alcohol-related harm, risky use, and high consumption. Finally, the study contributes to a limited but growing literature on the intersection of pricing policy and health equity.

Nonetheless, several limitations must be acknowledged. Alcohol use and harms were self-reported and subject to recall and social desirability bias. The cross-sectional design precludes causal inference. Sample sizes were small for some subgroups, particularly Indigenous respondents, which limited precision and may have inflated effect estimates when statistically significant results were detected (Button et al., 2013). Intersectional analyses (i.e., studying subgroups with multiple vulnerability factors) could not be conducted due to limited sample size. Our measure of PPSD does not capture temporal or promotional price variation and collapsing individual-level price distributions to a weighted mean inevitably loses information about the full range of prices paid. Additionally, our price data derive exclusively from off-premise retail sources; on-premise drinks were assigned equivalent off-premise prices, which would bias associations toward the null. However, the majority of respondents reported exclusively or predominantly off-premise consumption, suggesting this is unlikely to materially affect the primary findings. We also note that the specific products consumed on-premise cannot be identified, and the ratio of on- to off-premise consumption may not be consistent across product price points. We also assumed that prices observed in government owned stores were representative of those in private retailers; this approach is most valid in BC’s quasi-monopoly retail environment and would require additional validation before application in liberalised markets with greater within-product price variation. Finally, although a comprehensive set of covariates were adjusted for, residual confounding remains possible.

### Conclusion

This study uses a novel method that demonstrates lower alcohol prices are closely related to risky alcohol use and harms. By linking product-specific consumption data with provincial retail prices, we found that individuals who consumed cheaper alcohol had higher odds of causing alcohol-related harm to oneself or to others and scoring highly on the AUDIT.. These associations were strongest among socioeconomically disadvantaged and Indigenous respondents, making this the first study to estimate these affordability–risk relationships in this way directly among structurally vulnerable groups.

By providing individual-level price per standard drink estimates across population subgroups, this study fills a major evidence gap and offers granular data that can inform future modelling. These affordability profiles can be integrated into simulation and policy models to examine how changes in pricing, taxation, or market structures may influence consumption and harm across different groups.

Further work using longitudinal data, larger subgroup samples, and product-level modelling frameworks will be essential for identifying the alcohol products most linked to harm and for projecting how affordability-focused interventions may reduce inequities in alcohol-related outcomes.

## Data Availability

The survey data used in this study are not publicly available due to participant confidentiality and ethics restrictions. The British Columbia Liquor Distribution Branch (BCLDB) retail price data used for product linkage are available upon request from the BCLDB.

## Primary funding

This research was funded by the B.C. Ministry of Health Seed Grant Program.

## Conflict of interest

None to declare.

## Supplementary Material

### Appendix A. A comparison of the analytic sample versus eligible non-completers

**Table A1.** Comparison of key sociodemographic characteristics between the analytic sample and eligible non-completers.

| Variable | Analytic Sample (SD) | Non-Completers (SD) | p-value |
| --- | --- | --- | --- |
| N | 1,221 | 570 |  |
| Age | 54.70 (17.54) | 54.50 (18.84) | .829 |
| Sex |  |  | **.021** |
| Men | 58.89% | 52.98% |  |
| Women | 41.11% | 47.02% |  |
| Ethnicity |  |  | .433 |
| Arab or Middle Eastern | 1.15% | 0.35% |  |
| Asian | 18.47% | 13.49% |  |
| Black | 0.90% | 0.69% |  |
| Hispanic or Latin | 1.23% | 1.39% |  |
| Indigenous | 2.46% | 2.42% |  |
| Other | 0.66% | 0.35% |  |
| White | 72.74% | 77.85% |  |
| Prefer not to answer | 2.38% | 3.46% |  |
| Education |  |  | .469 |
| Some high school | 1.39% | 2.07% |  |
| High school or GED equivalent | 13.31% | 14.14% |  |
| College diploma or equivalent | 33.39% | 34.14% |  |
| Bachelors | 31.99% | 31.03% |  |
| Masters | 13.70% | 14.83% |  |
| Doctoral | 1.89% | 3.45% |  |
| Prefer not to answer | 0.33% | 0.34% |  |
| Income |  |  | .053 |
| < \$30,000 | 7.29% | 7.69% | |
| \$30,000-\$59,999 | 19.08% | 16.78% | |
| \$60,000-\$99,999 | 28.17% | 25.17% | |
| \$100,000-\$124,999 | 15.64% | 12.24% | |
| \$125,000-\$199,999 | 16.63% | 17.48% | |
| \$200,000 or more | 6.47% | 10.84% | |
| Prefer not to answer | 6.72% | 9.79% |  |

*Note.* All statistics are unweighted. p-values are from independent samples t-tests for continuous variables and chi-square tests for categorical variables, with Fisher's exact test used for ethnicity and education due to small expected cell counts in some categories.

### Appendix B. Product-selection module of survey

We captured product-specific alcohol consumption over a seven-day recall window using a branched module. Figure B1 shows the category and subtype screeners that established which beverage categories a respondent consumed at least once in the past seven days and, for wine and spirits, which subtypes were relevant. Selections here determined which product list were shown next to reduce burden.

**Figure B1.**
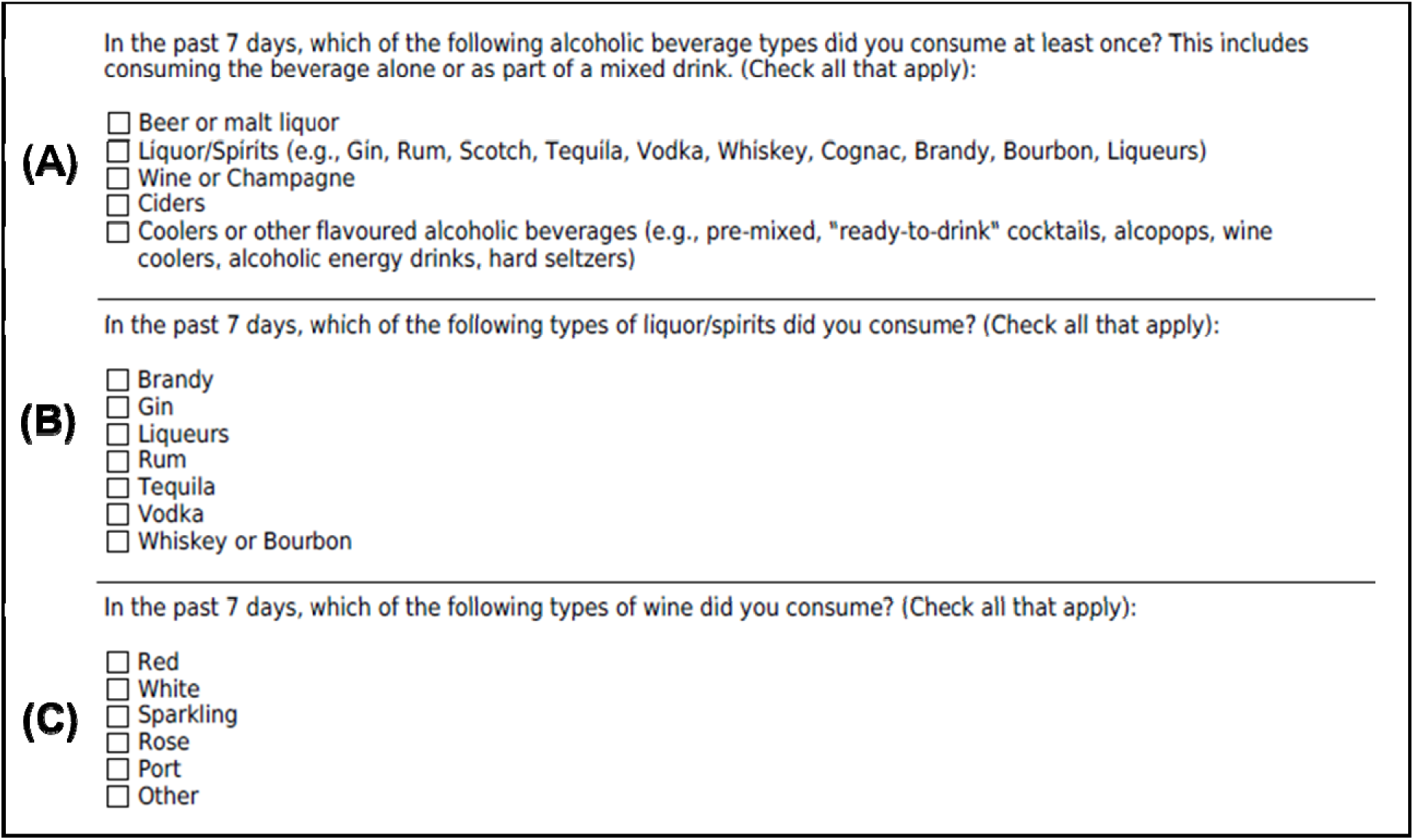
Beverage category and subtype screeners for the seven-day recall. *Note.* Panel A asks respondents to mark all beverage categories consumed at least once in the past seven days, including beverages taken as part of mixed drinks. Panel B appears only if Liquor or Spirits i selected and captures spirits subtypes. Panel C appears only if Wine or Champagne is selected and captures wine subtypes. Choices in Figure 1 determine which product selectors are shown subsequently.

**Figure B2** shows an example of the brand-to-product dropdowns used within each selection pathway. Respondents first chose the brand(s) consumed, then selected specific product(s) within brand. For every selected product, they entered a seven-day quantity using a numeric field. If binge drinking (defined as consuming five or more drinks on one occasion for men or four or more drinks on one occasion for women) was previously endorsed, a product-specific binge drinking question followed. Multi-select was enabled at both brand and product levels so respondents could report multiple item within a category.

**Figure B2.**
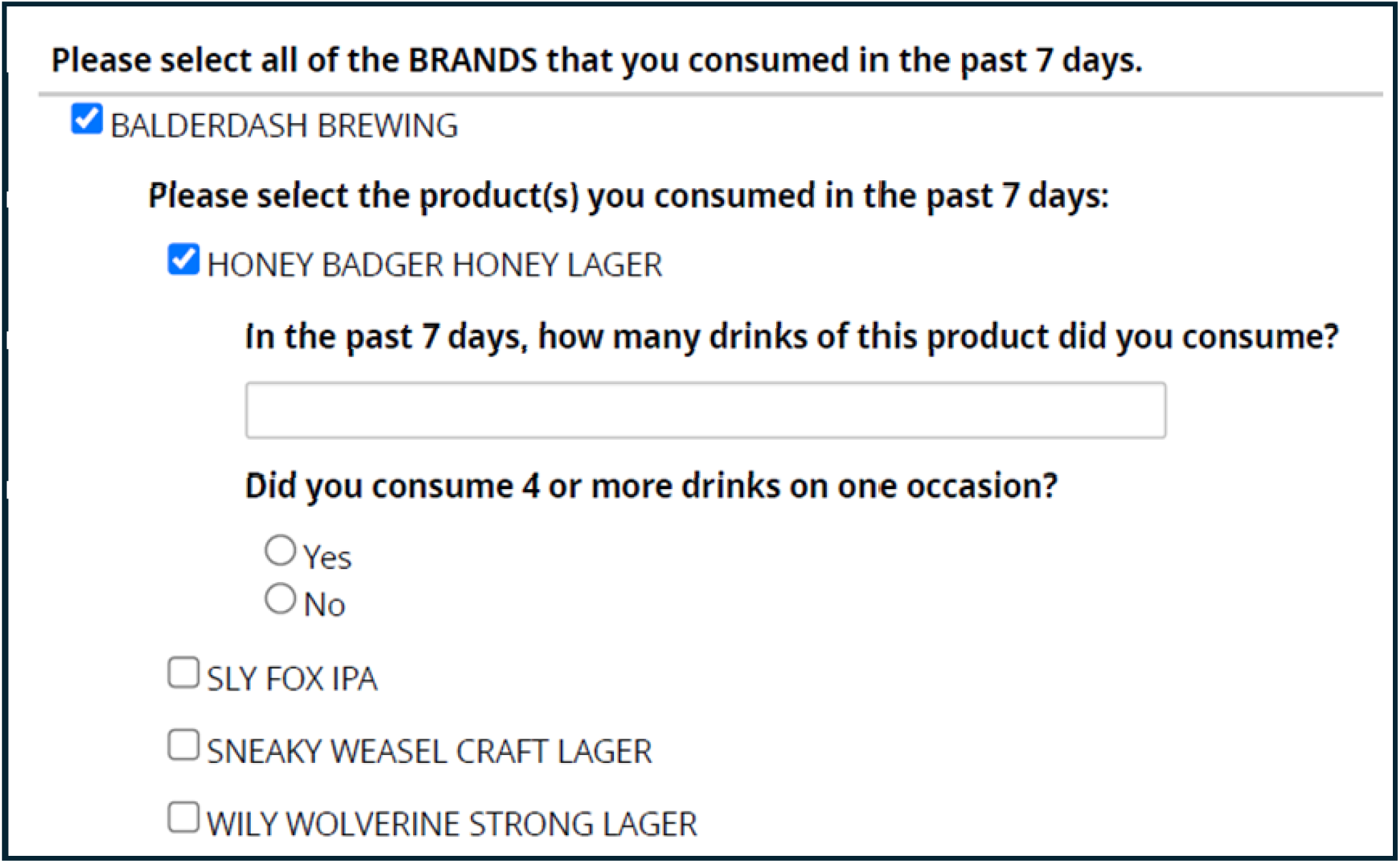
Brand-to-product dropdowns and alcohol consumption prompts. *Note.* Example display for a female respondent who previously endorsed binge drinking, showing a report of consuming Honey Badger Honey Lager from Balderdash Brewing in the past seven-days.

### Appendix C. On-premise consumption

**Figure C1.**
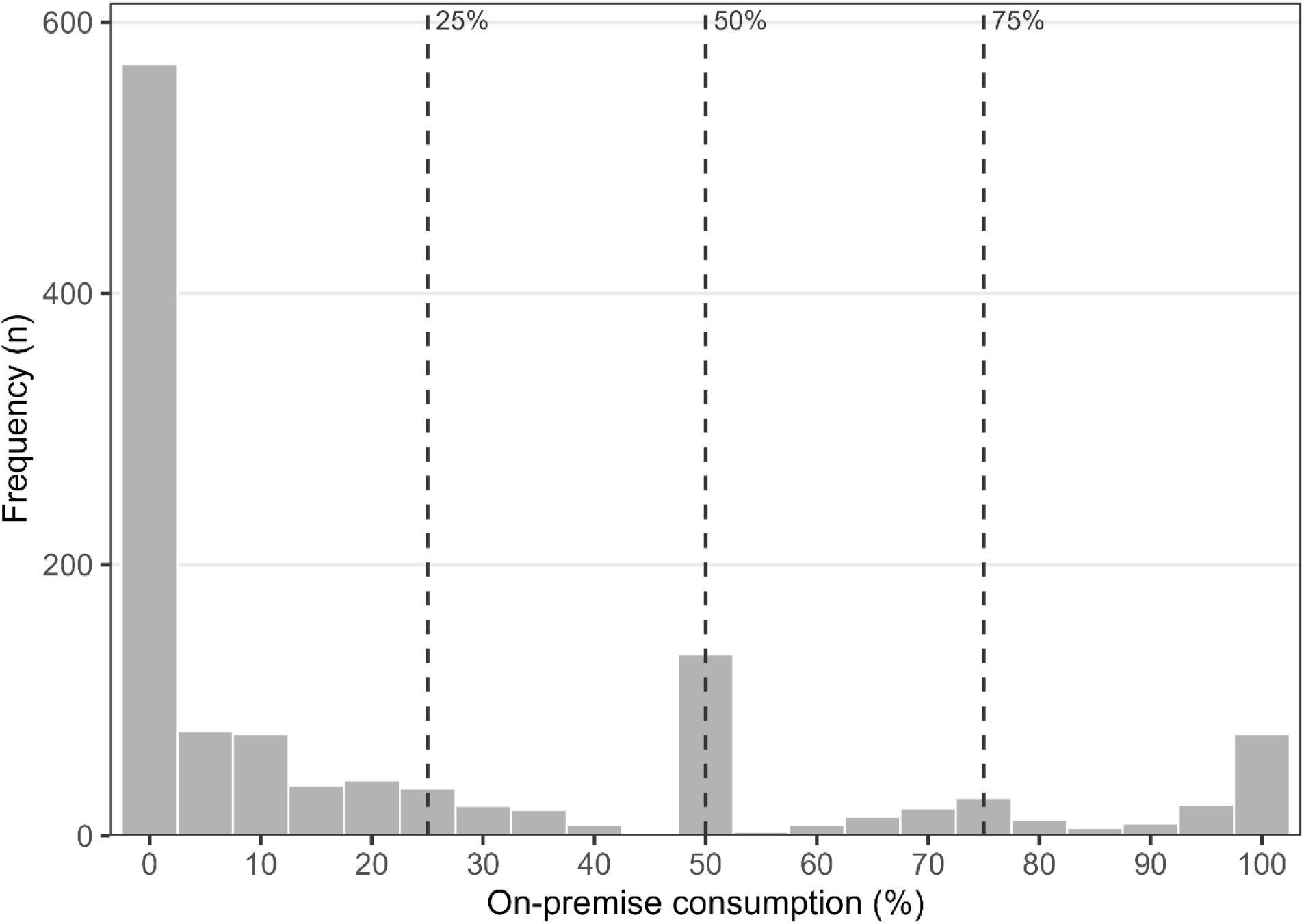
Distribution of self-reported on-premise alcohol consumption (e.g., in bars, restaurants, clubs) among past-week drinkers in British Columbia (*n* = 1,217). Dashed vertical lines indicate 25%, 50%, and 75% thresholds. Frequencies are unweighted counts.

## Notes

### Competing Interest Statement

The authors have declared no competing interest.

### Funding Statement

This research was funded by the B.C. Ministry of Health Seed Grant Program. No third party played any role in study design, data collection, analysis, interpretation, or manuscript preparation. The authors have no financial relationships with commercial entities that have an interest in the subject matter of this manuscript.

### Author Declarations

The University of Victoria Human Research Ethnics Board gave ethical approval for this work(#24-0235).

### Summary of Updates

This version of the manuscript has been revised to correct data errors and improve clarity. Errors in Table 1 and a coding issue affecting subgroup estimates in Table 3 have been corrected; these changes do not affect the overall conclusions. The framing of the statistical analyses has been clarified to emphasise that price per standard drink reflects exposure rather than a direct causal effect. The Introduction, Methods, and Discussion have been revised for clarity, including additional detail on data linkage, on-premise consumption, and sample representativeness. Minor edits were made throughout, and an additional co-author has been included.

